# How do primary care providers perceive their role at ensuring opioid safety? A qualitative exploration from those on the front lines

**DOI:** 10.1101/2022.04.22.22273055

**Authors:** Chris Gillette, Sarah Garvick, Edward Hak-Sing Ip, Robert Hurley, Julienne Kirk, Sonia Crandall

## Abstract

**Introduction:** Prescribing naloxone is recommended by the Centers for Disease Control and Prevention to reduce the risk of death from an opioid overdose. Naloxone is rarely prescribed, even when indicated; improving our understanding of how primary care providers (PCP) perceive their role in naloxone prescribing is essential to increase opioid medication safety. The objectives of this study were to: (1) describe how PCPs perceive their role in prescribing naloxone for patients who are at high risk of an overdose and (2) describe PCP-reported barriers and facilitators of naloxone prescribing.

**Methods:** Currently practicing providers completed semi-structured interviews, based on Theory of Planned Behavior, to understand their attitudes toward naloxone, their perceived role in naloxone prescribing, and facilitators/barriers to prescribing naloxone.

**Results:** Eleven interviews were conducted with physicians (n=2), physician assistants (n=8), and a nurse practitioner (n=1). Providers held generally positive attitudes toward naloxone as a ‘rescue’ medication. Negative attitudes toward naloxone include the perception of facilitating risky opioid use. Providers suggested that whomever prescribes the opioid pain medication should be primarily responsible for prescribing naloxone. Providers noted that stigma may prevent them from discussing naloxone during clinic visits. Increasing visit time and receiving support/education from organizational and professional society leadership were identified as important facilitators of naloxone prescribing.

**Conclusions:** While providers were aware of what naloxone was used for, there was reticence in discussing this medication with patients. Providers reported that whomever prescribes a pain medication should be primarily responsible for ensuring medication safety. If primary care organizations would like to improve opioid medication safety, ensuring that providers feel supported and receive needed education are essential.

## Introduction

The United States of America (US) is continuing to experience an epidemic of drug overdose deaths that is unmatched anywhere else in the world. The Centers for Disease Control and Prevention (CDC) recently reported that, in 2020, more than 93,000 Americans died from a drug overdose, an increase of almost 30% from 2019 and the highest number ever recorded in one year.^1^ As a result, the CDC is continuing to recommend naloxone to prevent opioid-related overdose deaths and an expansion of ongoing prevention and emergency response options.^2^ In addition to the staggering loss of life from drug overdoses from prescription and illicit opioids in the US, drug overdoses also impose a substantial economic cost. Using 2013 costs, the total economic burden in the US from prescription opioid overdose and dependence was $78.5 billion; it is, possible, due to increases in overdose death rates and inclusion of illicit drug overdoses, the economic cost from overdose and dependence could be well over $100 billion per year in 2021.^3^

Naloxone is an opioid antagonist that is used to reverse the effects of an acute opioid overdose.^4^ Naloxone works by binding to multiple opioid receptor sites and blocks the effects of other opioids. In the absence of an opioid, naloxone does not produce any pharmacological effect.^4^ Naloxone is not recommended as a treatment for opioid use disorder (OUD), but is recommended for patients and loved ones to keep close by, in case of an emergency. In the 2016 CDC opioid prescribing guidelines, naloxone was recommended to be prescribed for all patients who are at high-risk of experiencing an opioid overdose.^5^ Those who are at high-risk of an opioid overdose are those with a total daily morphine milliequivalent (MME) dose at or above 50MME, concurrent benzodiazepine and opioid medication prescriptions, and/or a previous history of substance abuse.^5^ A recent CDC Morbidity and Mortality Weekly Report (MMWR), however, established that only about 1 in every 100 patients with high-dose MME prescriptions had naloxone co-dispensed.^6^ It is also estimated that anywhere from 9%-25% of persons who use opioids also concurrently use benzodiazepines.^7-9^ Unfortunately, medication reconciliation problems are well-documented, including both time and resources, showing that during busy clinical practice, providers may be under-informed about all the medications and supplements their patients are taking, increasing the risk for opioid-related adverse drug events, including mortality.^10^ Since research has shown that both opioid days supply and concurrent use of opioid and benzodiazepines is increasing, it is imperative to close the gap between what is known to be effective and safe (e.g., prescribing naloxone) and what is currently done to ensure that patients are not harmed from their medication.^11-12^

A potential reason for such a low rate of naloxone dispensing is that many providers may hold negative perceptions about individuals who have a history of a substance use disorder.^13-14^ Provider stigma against these patients may negatively affect willingness to prescribe naloxone.^14^ Another recent study found that primary care providers and clinic staff reported substantial knowledge gaps regarding how to prescribe naloxone and who should get a prescription, showing that current efforts from healthcare organizations, such as the CDC, have not been effective and may have been counterproductive.^15^ One recent study conducted in a large, multi-state community health center network found that opioid prescriptions have dropped precipitously, which has been corroborated nationally by the CDC.^11,16^ The quick decrease in opioid prescribing may counterfactually increase the need for naloxone because patients who receive long-term opioids who have had prescriptions discontinued by their providers may seek relief from illicit opioids.^17^ A report from North Carolina found that one of the most common complaints the medical board receives from patients has been that their providers have suddenly stopped prescribing their opioids without further resources, which may help explain the continued increase in increasing mortality from illicit opioids while opioid prescribing has decreased.^17^

A recent study found that primary care physicians expressed the lowest support (52.6% supported) for providing naloxone to friends and family members of people using prescription opioid pain medication amongst a range of policies to reduce prescription OUD.^13^ In fact, support for this policy was markedly lower than arresting and prosecuting physicians who provide pain medications without an appropriate indication (63% supported). This low level of support for naloxone with the added societal stigma may be one reason why naloxone is rarely prescribed, even though current guidelines recommend it.^5-6^ Improving our understanding of how PCPs perceive their role in opioid treatment and naloxone prescribing is essential for policymakers and medical educators to design effective interventions if they want to increase opioid medication safety. Therefore, the objectives of this qualitative study were to: (1) describe how PCPs perceive their role in prescribing naloxone for patients who are at high risk of an overdose and (2) describe PCP-reported barriers and facilitators of naloxone prescribing.

## Methods

### Participants and Procedure

The Wake Forest School of Medicine Institutional Review Board (IRB) approved this study. The authors used Standards for Reporting Qualitative Research (SRQR) to guide the reporting of this study.^18^ A convenience sample of providers from an institutional listserv and the researchers’ personal contacts completed semi-structured interviews. Providers were eligible to participate in the study if they: (1) were at least 18 years of age; and (2) an actively practicing physician (MD/DO), physician assistant (PA), or nurse practitioner (NP) in a primary care setting. We defined primary care settings as: (1) family medicine clinics, (2) general internal medicine clinics, and (3) urgent care clinics. We based the interview guide on the Theory of Planned Behavior (TPB) (**Figure 1)**.^19^ The TPB states that the best predictor of a person performing a behavior is behavioral intention. Attitudes toward the behavior, peer referents, and control beliefs all influence intention, and therefore, behavior. We designed the interviews to explore attitudes toward prescribing naloxone and educating patients and their families about how to use it, their perception of how other providers educate about naloxone, and what would make it easier/harder to educate patients and their loved ones about naloxone. The TPB is frequently used as a theoretical framework in implementation science research in a variety of settings and health conditions.^20^ We collected and analyzed data from September 2020 through April 2021.

**Figure 1:**
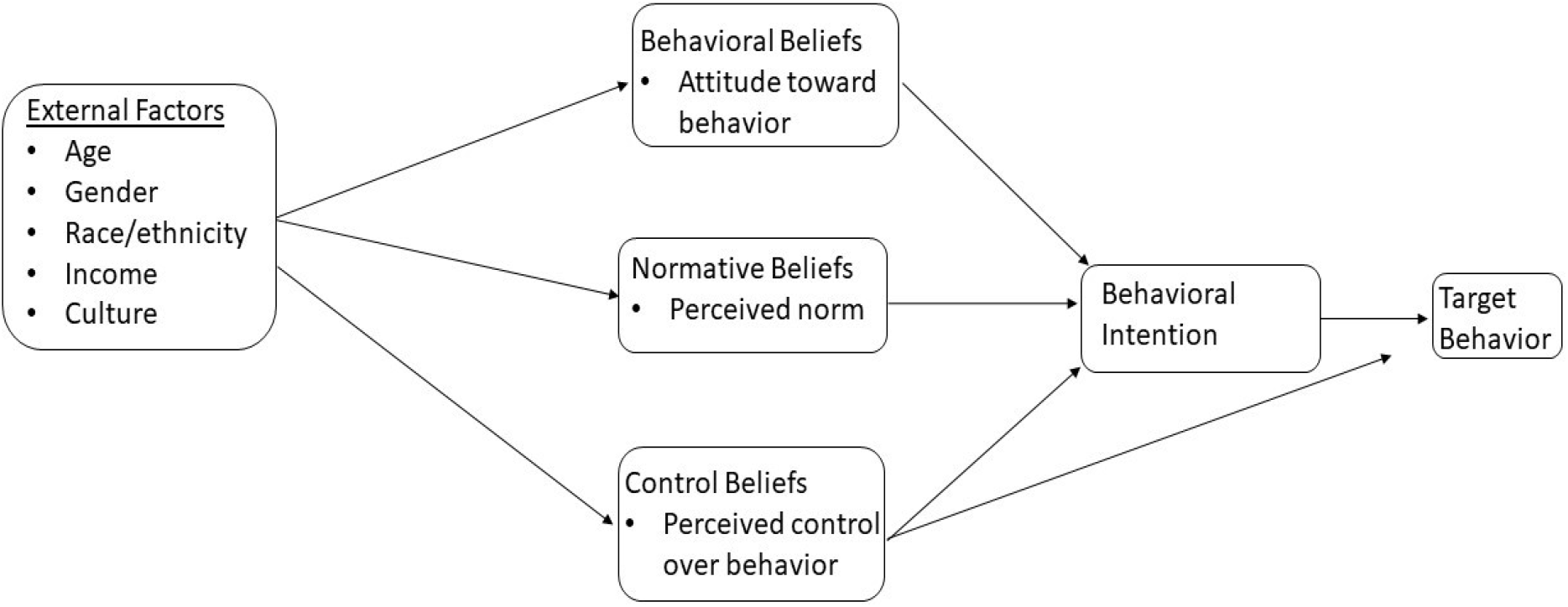
Theory of Planned Behavior Theoretical Model.

Two researchers conducted the interviews (CG and SG). The first author (CG) is an experienced pharmaceutical outcomes researcher with a background in both qualitative and quantitative methodology. The PA researcher (SG) is a primary care PA who currently practices in a rural area. The first (CG) and senior authors (SC) are experienced researchers and collaboratively trained the PA author in interview techniques for qualitative research as well as the qualitative analyses described below.

A regional area health education center (AHEC) sent emails with links describing the study to healthcare providers. The chosen AHEC consists of 17 counties within North Carolina who, according to the North Carolina Department of Health and Human Services, have some of the highest need for naloxone.^21^ Providers were instructed to input their contact information if they were interested in participating in the study. The interviewers contacted participants to further explain the study, answer their questions, and set up a mutually agreeable time for the full interview. We recruited eleven (n=11) primary care providers: 2 physicians, 8 PAs, and 1 NP. All but one (n=10) identified as non-Hispanic white, three (n=3) identified as male gender, with an average age of 34 years.

### Interviews

The interviews consisted of 15-45 minute semi-structured interviews that explored participants’ experiences prescribing and educating patients about naloxone. Examples of questions that were asked included (but are not limited to): (1) How often do you personally prescribe naloxone and how do you decide when you want to prescribe it?; (2) what do you see as the advantages of educating patients and/or their families about how to use naloxone?; (3) please describe any factors that would make it easy for you to educate patients or their families about how to use naloxone.

### Data Analysis

Each interview was transcribed to ensure coding accuracy. Transcribing audio interviews into text facilitates data analysis. We analyzed data using constructivist grounded theory, which is an inductive approach that allows theory generation and construction. Grounded theory is appropriate to use when the phenomenon under study has not been previously studied. We are unaware of any research that has investigated the influence of PCP attitudes, norm referents, and control beliefs when educating patients about how to use naloxone. To ensure rigor and dimensionality in interpretation, two authors (CG and SG) conducted the data analysis using DeDoose, which is a cloud-based software that allows for multiple researchers to remotely analyze textual data.

## Results

### Attitudes toward Naloxone Education

The providers reported generally positive attitudes toward naloxone and naloxone education, stating that the primary benefit of naloxone is that it saves lives in case of an opioid overdose. Most providers, however, stated they do not routinely prescribe naloxone to patients they believe to be at high risk of opioid overdose. Providers also stated that the benefits of educating patients about naloxone is that conversation allows them to combat misinformation and empower patients and their families regarding saving lives. The quotes below exemplifies the general attitude toward naloxone and naloxone patient education:

> *Provider 7: “I think it has to start with some leveling the playing field and getting on the same page, and ground rules and understanding what it’s for and the appropriate use and utility of it. But I think the main advantage is the avoidance and the turn of events of the outcome, that being death of an opioid overdose, actually can lead to. So, in my opinion, the biggest advantage is being able to have this medication more readily accessible and prescribed maybe a little bit more routinely by providers or those with the prescribing privileges, is ultimately detouring away from the outcome, that being death in the setting of an overdose*.*”*
>
> *Provider 6: “But even when I give opioids…I always tell the patient, ‘This could lead to addiction, you’ve seen those people, you’ve heard the stories about those people’…So I really think that patient education is the best way to go. And if you take your time to explain to them, what happens when you take opioids and, why it leads to constipation, why people feel groggy and sedates [sic]. I think that really helps them understand what’s going on. And when you give patients knowledge, they’re now empowered about their health*.*”*

The providers also reported naloxone is a key component to ensuring safety with opioid pain medications. The providers who spoke of opioid safety compared naloxone to a ‘rescue medication’ similar to albuterol for patients who have acute asthma symptoms or an Epi-Pen for those with allergies. The quotes below summarizes how providers thought of naloxone in the context of opioid safety:

> *Provider 9: “ …I think it’s a very important thing to probably have a conversation about it and I think we should probably do it more often than we do. Really, and you can read anywhere, there’s no truly safe dose of a narcotic. It doesn’t matter how little someone gets. There’s always an opportunity of having a bad outcome…. And it’s just as important to talk about how do we mitigate those risks? And I think that naloxone is an answer. It’s not the answer, or certainly not the only answer, but I think it probably does play an important part in the discussion that we probably should [be]having [sic]*.*”*
>
> *Provider 4: “Yeah, I mean it’s life-saving. It’s kind of, in my mind, an equivalent to an EpiPen, but in a different circumstance. If someone’s throat was closing up because of an anaphylactic reaction, we would want them to have Epi [sic] to save their life. I think it’s a similar situation with [naloxone], except that it’s complicated because of the social context. But it’s similar in my mind, in that it’s a rescue*.*”*

### Disadvantages of Prescribing Naloxone and Patient Education

While providers were generally supportive of naloxone and educating patients about naloxone, some providers did express reservations about prescribing naloxone. The primary concern about naloxone was the perception that prescribing naloxone facilitates risky opioid use by removing patient concerns about dying from an opioid overdose. Providers also reported that educating patients about naloxone would negatively impact the provider-patient relationship because of two main issues: (1) patients believe that opioid pain medications are safe, especially those who may have long-term use of opioids, and (2) offending patients by making them believe they are addicts. Providers noted that there is a particular social stigma that negatively impacts the perception about long-term opioid use that they are reluctant to address with patients. The following quotes summarize these disadvantages:

> *Provider 8: “…I think the only disadvantage is that when I spoke of earlier, that ethical political slippery slope argument that if I’m giving you this medicine, I’m essentially giving you free license to misuse opioids or use them in a way you weren’t supposed to or should and you’ve got an artificial safety net. That’s really the only potential downside that I personally see or have heard folks talking about*.*”*
>
> *Provider 4: “Because I think people who may be annoyed or they may be like, ‘Oh, I don’t want to have to do this’, or, ‘This doesn’t apply to me”, or, ‘This PA doesn’t know what she’s talking about. My primary care [provider] has been prescribing this medicine for 20 years”*.

### Who Is Responsible for Educating Patients about Naloxone?

Providers reported that pain specialists were the most likely type of provider who would prescribe naloxone to patients who were at high risk, followed by PCPs. Providers also mentioned other providers who may prescribe naloxone, including psychiatric and behavioral health specialists. The below quote exemplifies the provider responses:

> *Provider 8: It’s probably going to be the pain management specialist. Folks who are working in those clinics and utilizing opioids on a regular basis. I think those are the folks where you would find that education going on most commonly and find the folks who have figured out how to do it the best would be…because again, so many providers are not prescribing opioids compared to just five years ago. …I think that’s appropriate, but I think there’s a general hesitancy and I think, so the folks who are doing it well are the ones who are doing it a lot and you’re going to find them mostly in pain management scenarios*.

### Barriers to Educating Patients about Naloxone

Providers mentioned multiple barriers that prevent them from routinely educating patients about naloxone, most of which was limited time during clinic visits. Providers also noted that stigma in general and in particular, provider stigma about patients who may be at high-risk for an opioid overdose contribute to them not educating patients about naloxone. The below quote highlights the stigma barrier that providers face about naloxone and substance use disorders:

> *Provider 11: “And too, the patient, I think from an education standpoint, might not realize that there’s something out there that they can have, or they can have a prescription for, or they don’t know to ask about it. Or maybe they want to ask about it, but there, again, is a stigma associated with it and they don’t want to ask about it. Again, that’s my opinion and it’s certainly not backed up by any research that I’ve done or anything like that. But I think that of the people that I’ve prescribed it for, I think a lot of them could probably use it, but I think, equally, that they probably would not ask for it*.*”*
>
> *Provider 8: “So whether you’re a person who doesn’t prescribe them or someone who works in a pain clinic [and] prescribes them regularly, it’s an uncomfortable topic. It’s so personal to speak about it because there’s such a stigma around opioid use and particularly opioid misuse. I think it’s too easy to label people as making bad choices and not necessarily label them correctly with a condition like diabetes or pneumonia that’s really a medical one. I think there are a lot of layers to it and it’s not real crystal clear and when you get out of medicine and into society in general, I think it’s even more clouded*.*”*

### Methods to Improve Naloxone Patient Education

While providers were generally supportive of prescribing naloxone, few reported prescribing naloxone or educating patients about how to use naloxone. To improve naloxone prescribing, providers noted that support from organizational leadership and professional societies was important to increase the amount of providers who are educating patients about naloxone. The quote below exemplifies the provider responses regarding organization and professional society support to help them educate patients about naloxone:

> *Provider 10: “I think anytime a practice*… *has consensus around a certain practice…then that makes it easy…*.*‘This is our stance as a group, and this is something that we feel is important’…*.*And then if you have questions, if everybody’s been given the same information, then you could ask any of your colleagues if a question came up, versus feeling like, ‘Well, I’m the only one who is really doing this. I don’t really have people to ask*.*’ That could be a major deterrent, or just an obstacle*.*”*
>
> *Provider 4: “Probably the American Academy of Family Physicians, because they’re who I go to for most information. That’s kind of my wheelhouse. So if they’re pushing it, then it makes it seem like something that I can do and that I should be doing. Because especially in primary care, we always walk a fine line between, of course, wanting to do everything that we can for our patients, but then also not wanting to cross into other specialties where there are specialists who are managing certain conditions and undermining them or acting as though we’re better at managing something than they are. So, I think that if they [AAFP] were saying, ‘Even if your patient is prescribed opioids by someone else, you should be prescribing this [naloxone]at least once every six months or once a year, depending on when it expires’, then I think that would be something that I would really listen to and would kind of influence me*.*”*

Providers also identified educational needs they felt would increase their comfort and ability to properly educate patients about naloxone and how to use it. Notable educational needs that providers identified were: (a) naloxone adverse effects and (b) costs/third-party coverage. Interviewees also noted that having standardized informational brochures that used patient friendly language and/or a naloxone demonstrator would help facilitate provider-patient conversations regarding naloxone.

## Discussion

Naloxone has been shown to be safe and effective for patients who are experiencing an opioid overdose.^4^ Despite this, providers rarely prescribe and educate patients on how to use it.^6^ Our in-depth interviews with primary care providers show that while providers are generally supportive of naloxone, there are significant organizational and educational barriers that may prevent them from prescribing and educating patients about it. These barriers include not having enough information/education about naloxone, stigma, and worrying about facilitating riskier opioid use. Providers noted that organizational and professional support is needed if more frequent naloxone prescribing is desired.

The moral hazard concern about naloxone is not new even though there is little evidence to support it.^22-24^ In 2016, Coffin and colleagues found that co-prescribing naloxone to patients in primary care settings may decrease the occurrence of opioid adverse events and reduce Emergency Department (ED) visits compared to those who did not receive naloxone.^25^ Moreover, the authors did not find increased prescription opioid dosages among those who received a naloxone prescription compared to those who did not. A more recent study found that long-term chronic pain patients who received naloxone along with their opioids had significantly fewer opioid-associated ED visits than patients who did not receive naloxone, showing that the moral hazard concern is unfounded.^26^

Our finding that providers are hesitant to prescribe and educate patients about naloxone due to stigma has also been observed in other studies of provider concerns about naloxone prescribing.^27-28^ Providers in this study feared that discussing naloxone may make the patient feel that they were being labeled as an ‘addict’. Previous research has found that patients are reluctant to bring up the need for naloxone because they fear that doing so may cause a provider to believe they are misusing their pain medication.^29^ This fear from providers may be unfounded because prior research providers in San Francisco felt that naloxone improved the provider-patient relationship and this conversation made them think about their opioid prescribing more carefully.^30^ The study from Behar and colleagues was conducted in primary care clinics who had initiated a formal naloxone training and co-prescribing intervention whereas the clinics in our study had not received any such intervention, which may help explain some of the discrepancy.. Providers who would like to discuss naloxone with their patients should do so using empowering and non-judgmental language, framing the discussion as a “worst-case scenario”. Patients also desire more information and training on how to recognize when to use naloxone and how to use it correctly.^29^

Another important result from our study is these PCPs believed they have a role in naloxone prescribing, however, most providers stated that pain medicine and psychiatry/behavioral medicine should be primarily responsible for educating patients about how to safely use their opioid pain medications. This is an important and disappointing finding. The American Academy of Family Physicians (AAFP) defines a primary care physician and provider as: “…a specialist in family medicine, general internal medicine…who provides definitive care to the undifferentiated patient at the point of first contact, and takes continuing responsibility for providing the patient’s comprehensive care…Primary care physicians advocate for the patient in coordinating the use of the entire health care system to benefit the patient.”.^31^ Thus, ensuring medication safety for their patients is a key role for PCPs, regardless of who prescribed their patients’ medications. PCPs can also work with local community pharmacists to help coordinate and ensure medication safety by educating patients about the importance of naloxone.^32^ Medical educators can play an important role in safeguarding opioid medication safety by integrating this education throughout their curriculum to give future providers the tools necessary to reduce stigma and ensure patients receive proper access to needed medications.^33^

Our study has limitations that affect generalizability of its findings. The study was conducted among providers in one state in the southeast United States and may not be generalizable to other areas. Our sample size, similar to other qualitative studies, is small; this limitation may be mitigated, however, because our findings have also been shown in previous studies with larger samples. Further, as Guest and colleagues point out, qualitative data saturation is an elastic concept that is based on numerous assumptions, one of which is homogeneity of the sample.^34^ The purpose of our project was to better understand healthcare providers’ attitudes toward educating patients about naloxone. Our goal was not to compare the experiences of physicians, PAs, and NPs in this phenomenon, so we included them all together as healthcare providers.

Further, we undertook several steps to ensure that participants were relatively homogenous, even though they were from different professions. First, we restricted the sample to PCPs, regardless of profession. This would ensure that these providers have experience treating a wide range of conditions for which high-risk opioid users would be a part. Second, participants were taken from one region in the United States to further ensure homogeneity within the sample. Third, the objectives of our study were fairly narrow, limiting the project to provider-patient communication about one specific drug. Even though the content of the discussions tended to also include other medications (e.g., opioids and benzodiazepines), the discussions were fairly limited to naloxone. Providers may have had additional reasons why they were hesitant to prescribe and educate patients about naloxone they did not reveal to the interviewers and they may have altered their true feelings about naloxone patient education due to social desirability bias.

While our study has limitations, our study is a valuable addition to the literature because it is the first that we are aware of to use a theoretical framework to approach provider-patient communication about naloxone and to explore this communication in the southeast US. We found little variance in the behavioral beliefs about naloxone because almost every provider had generally positive attitudes toward educating patients about naloxone. The TPB’s constructs of normative and control beliefs, however, may hold promise as a way to improve provider-patient communication about naloxone. For instance, these providers also reported that the complex relationship between PCPs and specialists may hinder their willingness to discuss naloxone with their patients, reflecting the social norm construct in the TPB. These providers also reported reduced behavioral control because they feel under-informed about naloxone, especially when it comes to adverse effects. Future educational interventions to increase naloxone prescribing may need to focus on reducing provider-perceived stigma, increasing focus on medication attributes, such as adverse effects, and how to effectively communicate about naloxone to reduce patient-perceived stigma.

## Data Availability

All data produced in the present study are available upon reasonable request to the authors.

## Acknowledgements

The project described was supported by the National Center for Advancing Translational Sciences (NCATS), National Institutes of Health, through Grant Award Number UL1TR001420. The content is solely the responsibility of the authors and does not necessarily represent the official views of the NIH. The authors wish to thank the Northwest Area Health Education Center (NW AHEC) for their role facilitating the provider-patient interviews throughout the study period and Ms. Carol Hildebrandt for her role in ensuring this was a successful project.

